# Initiation and completion of treatment for latent tuberculosis infection in migrants globally: A systematic review and meta-analysis

**DOI:** 10.1101/2021.06.09.21258452

**Authors:** Kieran Rustage, Jessica Lobe, Sally E. Hayward, Kristina L Kristensen, Ioana Margineanu, Ymkje Stienstra, Delia Goletti, Dominik Zenner, Teymur Noori, Manish Pareek, Christina Greenaway, Jon S. Friedland, Laura B Nellums, Sally Hargreaves, on behalf of ESGITM and ESGMYC

## Abstract

**Background:** Latent Tuberculosis (LTBI) is one of the most prevalent infections globally and is key in development of active tuberculosis disease (TB). In many low-burden countries, LTBI is concentrated within migrant populations reflecting higher disease burden in some countries of origin; national programmes may consequently focus on screening and treating LTBI in migrants to prevent future TB cases. However, little is known about the extent to which migrants initiate treatment for LTBI when testing positive, and their treatment outcomes, which is urgently needed if we are to strengthen these programmes, improve migrant health, and meet TB elimination targets.

**Methods:** We did a systematic review and meta-analysis, following PRISMA guidelines and PROSPERO registered (CRD42019140338) to pool global data on LTBI initiation and completion amongst migrants (defined as foreign born), and secondary outcomes to explore the range of both personal and provider level factors associated with initiation and completion. We searched Embase, Medline and Global Health, and hand-searched grey literature (from Jan 1 2000 to Apr 21 2020). Inclusion criteria were primary research articles reporting on LTBI treatment initiation and/or completion amongst migrants; we excluded papers where data were not stratified by migrant status, or where the data related to outcomes prior to the year 2000. There were no geographical or language restrictions.

**Results:** 39 publications were included from 13 countries, with treatment initiation and completion data for 31,598 LTBI positive migrants. Overall, 69% (95% CI⍰=⍰51–84%; I^2^⍰=⍰99.62%) of these initiated treatment; 74% (95% CI⍰=⍰66–81%; I^2^⍰=⍰99.19%) of migrants who initiated treatment, completed it; among studies with data on the complete pathway from screening positive to completing treatment, 52% (95% CI⍰=⍰40–64%; I^2^⍰=⍰98.90%) successfully completed treatment. Meta-regression showed that LTBI programmes are improving, with more recent reported data (2010-2020) associated with better rates of treatment initiation and completion. European studies also appeared to have more successful outcomes than those in the Americas and Western Pacific WHO regions.

**Conclusions:** LTBI treatment initiation and completion amongst migrants have room for improvement. Though the data show improvements in the past decade, the delivery of these programmes will need further strengthening if we are to meet targets to eradicate TB in low-incidence countries. Greater focus will need to be placed on engaging migrants more effectively in the clinic and understanding the diverse barriers and facilitators to migrants initiating and completing treatment. Such efforts must be mindful of, and sensitive to the unique experiences individuals arriving in a new country.

**Funding:** This study was funded by the European Society of Clinical Microbiology and Infectious Diseases (ESCMID) through a joint ESCMID Study Group for Infections in Travellers and Migrants (ESGITM) and ESCMID Study Group for Mycobacterial Infections (ESGMYC) Study Group Research grant, the Rosetrees Trust (PhD studentship grant M775), the NIHR (NIHR Advanced Fellowship NIHR300072), and the Academy of Medical Sciences (SBF005\1111).

Panel: Research in Context

Evidence before this study
Latent tuberculosis infection (LTBI) is one of the most prevalent infections globally, affecting an estimated 25% of the population; re-activation of LTBI is a major driver of tuberculosis (TB) cases worldwide. In low-incidence TB countries, TB and LTBI are often disproportionately concentrated amongst foreign born individuals, with national programmes increasingly focusing on the diagnosis and treatment of LTBI in migrants and other high-risk groups to prevent future TB cases and meet global elimination targets. However, little is known about the success of these programmes in engaging migrants and ensuring treatment completion – a population who often face multiple barriers to accessing health care on arrival to the host country. Prior to this review we scoped the literature and found two relevant reviews on this topic (Sandgren et al., 2016 & Alsdurf et al., 2016) but which did not specifically focus on migrants and/or lacked formal meta-analyses, and one/both used earlier data pre 2000 that may be less relevant now to current policy. Other studies have reported on migrant-specific outcomes in LTBI programmes globally, but the focus is often on screening practices rather than outcomes and all evidence in this area has not yet been effectively consolidated.

Added value of this study
This is the first systematic review and meta-analysis specifically exploring LTBI treatment initiation and completion among migrant populations. We report LTBI treatment outcome data on 31,598 migrants from the year 2000 onwards within 13 low-incidence countries (<10 cases per 100,000). The research provides robust insights into the proportion of individuals initiating and completing treatment, using meta-regression to explore heterogeneity. The data show that between 2000-2020, 69% of migrants testing positive for LTBI initiated treatment, and of those starting treatment, approximately 74% completed it. Amongst studies capturing data on both initiation and completion, 52% of LTBI positive migrants successfully initiated and completed. The data also indicate higher initiation and completion in more recent years (2010-2020) with renewed focus on this approach to TB control, and a trend toward more positive outcomes amongst migrants in programmes in the WHO European region. The data show that multiple complex factors impact on treatment outcomes in migrants, including patient demographics and health systems. The evidence was ambivalent with some studies demonstrating positive and detrimental outcomes associated with foreign-born status.

Implications of all the available evidence
Delivery of LTBI programmes will need to be strengthened to improve outcomes in migrants and meet targets to eradicate TB in low-incidence countries. Greater focus will need to be placed on engaging migrants more effectively in the clinic, understanding the varied reasons for migrants’ declining treatment when testing positive, and ensuring treatment adherence using innovative approaches that are mindful of and sensitive to the unique experiences of this group on arrival to the host country.

## Introduction

Estimates indicate that 25% of the global population have Latent Tuberculosis Infection (LTBI);^1,2^ approximately 5-15% of those infected will develop active TB in their lifetime.^3,4^ Delivering effective LTBI screening and treatment programmes is increasingly being emphasised in the control of tuberculosis (TB), with the End TB strategy outlining expanded preventative treatment of people at higher risk of TB, and encouraging research that will improve the detection and treatment of LTBI.^5^ The first high-level meeting on tuberculosis in 2018 set a target to reach 30 million people with TB preventative treatment for LTBI between 2018-2022, including 20 million household contacts, and 4 million children under 5.^6^

In many high-income low-incidence TB countries (< 10 cases per 100,000)^7^ migrants experience a greater burden of TB relative to native-born populations.^8^ The majority of cases are likely due to re-activation of LTBI;^9^ the incidence of TB amongst migrants being greatest within the first 5 years of arrival driven by host factors (such as age and comorbid status) and socioeconomic factors (such as living conditions).^8,10^ The World Health Organisation (WHO) has published recommendations for low-incidence countries to consider systematic testing and treatment of migrants for LTBI.^11^ Consequently, the European Centre for Disease Prevention and Control (ECDC) has developed guidance for the programmatic management of LTBI within migrants (among other groups) in line with WHO recommendations.^12^

Increasingly, LTBI and TB screening programmes in low-incidence TB countries now include migrants from high-burden TB countries alongside other high-risk groups,^13^ where evidence indicates a greater risk of LTBI re-activation.^10,14^ Coinciding with this evolving focus, 6-9 month courses of Isoniazid therapy for LTBI are being increasingly replaced with shorter treatment regimens which often confer increased treatment completion and a reduction in complications and side effects for patients, which may have a positive impact on treatment completion rates going forward. These regimens include 3 months of weekly rifapentine (RPT) & isoniazid (INH), 3-4 months of isoniazid (INH) & rifampicin (RMP), 3-4 months of rifampicin (RMP) and 4 months of Rifapentine (RPT).^11,15^ Whilst availability and use of these regimens varies by country, it is plausible they could contribute to improved treatment completion amongst migrant patients.

Challenges and uncertainties exist regarding the current effectiveness of LTBI screening and treatment programmes involving migrants. A recent systematic review on infectious disease screening amongst migrants to Europe reveals such screening often focuses on LTBI, but suggests 54% of migrants with a positive LTBI test complete treatment.^16^ A further systematic review and meta-analysis analysing the entire LTBI care cascade for numerous populations groups (homeless, healthcare workers etc), but which included a minority of migrant patients, estimated that only 54.6% of migrants start treatment and 14.3% complete.^17^ Migrant specific factors, such as legal status was repeatedly highlighted as a risk factor for LFU when recommending, starting and completing LTBI treatment.^17^ The complexity of an individuals’ legal status can lead to fear and mistrust on the part of the patient where they have reason to believe engagement with health services can lead to detention and deportation, resulting in LFU. This can be further compounded by healthcare staff and vulnerable populations being unaware of rights to accessing health care, which can further present a barrier to treatment.^18^ Language and cultural barriers can also exist where health services have not adapted to meet the needs of diverse patient groups.^17,19^ More generally, clinicians recommended treatment may deviate from national guidelines due to clinician perception of the risk-benefit balance of treating LTBI.^17,19^ Whilst there are also general logistical barriers to accessing treatment (such as wait times) and side-effects which impact on all population groups at risk for LTBI.^17^Understanding the effectiveness of LTBI treatment programmes involving migrants, and further elucidating the factors underpinning this, is essential to contributing toward the reduction of LTBI and elimination of TB. Facilitators including having culturally sensitive services, and increasing patient involvement and ownership in delivering care, and ensuring high-quality service provider management aid screening; it would not be unreasonable to assume similar facilitators could be beneficial downstream during LTBI treatment.^16,18^ Fully understanding and addressing the facilitators and barriers to LTBI treatment initiation and completion will ultimately lead to the improvement of health outcomes for these individuals.

We therefore did a systematic review and meta-analysis to explore and assess available evidence on the initiation and completion of LTBI treatment amongst LTBI-positive migrants globally to better understand whether, and where efforts to improve current practices could be targeted.

## Methods

We did a systematic review and meta-analysis to explore both the initiation of treatment once a migrant had tested positive, and the completion of the full recommended course of LTBI treatment post-screening. We defined migrants as any foreign-born individual. Primary outcomes were treatment initiation, defined as the number of LTBI-positive migrants that successfully initiated LTBI treatment; and completion, defined as completion of the full recommended course of treatment according to the individual study specific parameters presented. Secondary outcomes were to explore and assess factors presented by the included studies associated with initiation and completion of LTBI treatment amongst migrants.

This research was carried out in line with the Preferred Reporting Items for Systematic Reviews and Meta-Analyses (PRISMA),^20^ and registered with PROSPERO (CRD42019140338).

### Search strategy and selection criteria

We included publications reporting primary data on LTBI treatment uptake and adherence in migrant populations. There were no geographic or language restrictions regarding the publications included, and we included both adult and paediatric populations. We excluded case reports and case series due to the limited number of individuals these study types would include. We also excluded publications with no primary data or discernible study design (e.g. comments, editorials, letters and reviews). We excluded studies with no clearly identifiable migrant population, or where the definition of a migrant conflicted with our own (for example defining migrant status on the basis of ancestry or ethnicity). We searched for publications from 2000 onwards, excluding those that reported on data from pre-2000, or where post-2000 data could not be disaggregated. We chose 2000 as cut off to better represent the rapidly evolving field in LTBI treatment in the past two decades, including contemporary treatments and global initiatives.

We searched the databases MEDLINE (1946 – 21/04/20), Embase (1974 – 21/04/20) and Global Health (1973 – 21/04/20) using a Boolean search strategy with keywords and relevant medical subheadings (MeSH) pertaining to three main themes: migrants, initiation & completion, and LTBI. Search terms were identified by consulting existing literature and experts in these areas. The search strategy is available in the appendices (appendix 1.) We explored the grey literature by hand searching conference proceedings and the websites of relevant NGOs and organisations (e.g. WHO). We identified additional publications through hand searching the bibliographies of publications included in the analysis, hand searching conference proceedings. We further compiled a list of experts in LTBI and emailed them formally requesting grey literature and publications they may be familiar with, or programmatic data they were at liberty to divulge.

Two reviewers duplicated the title and abstract screening and full-text screening (KR & JL), which was carried out using the web-based application Rayyan ^21^. The reasons for excluding studies during full-text screening were recorded (Fig 1.). Data was extracted by two reviewers (KR and JL). Where there were discrepancies in screening decision, or the extracted data between the primary reviewers, a third reviewer (LN or SH) mediated screening decisions and ensured the accuracy of extracted data.

**Figure 1.**
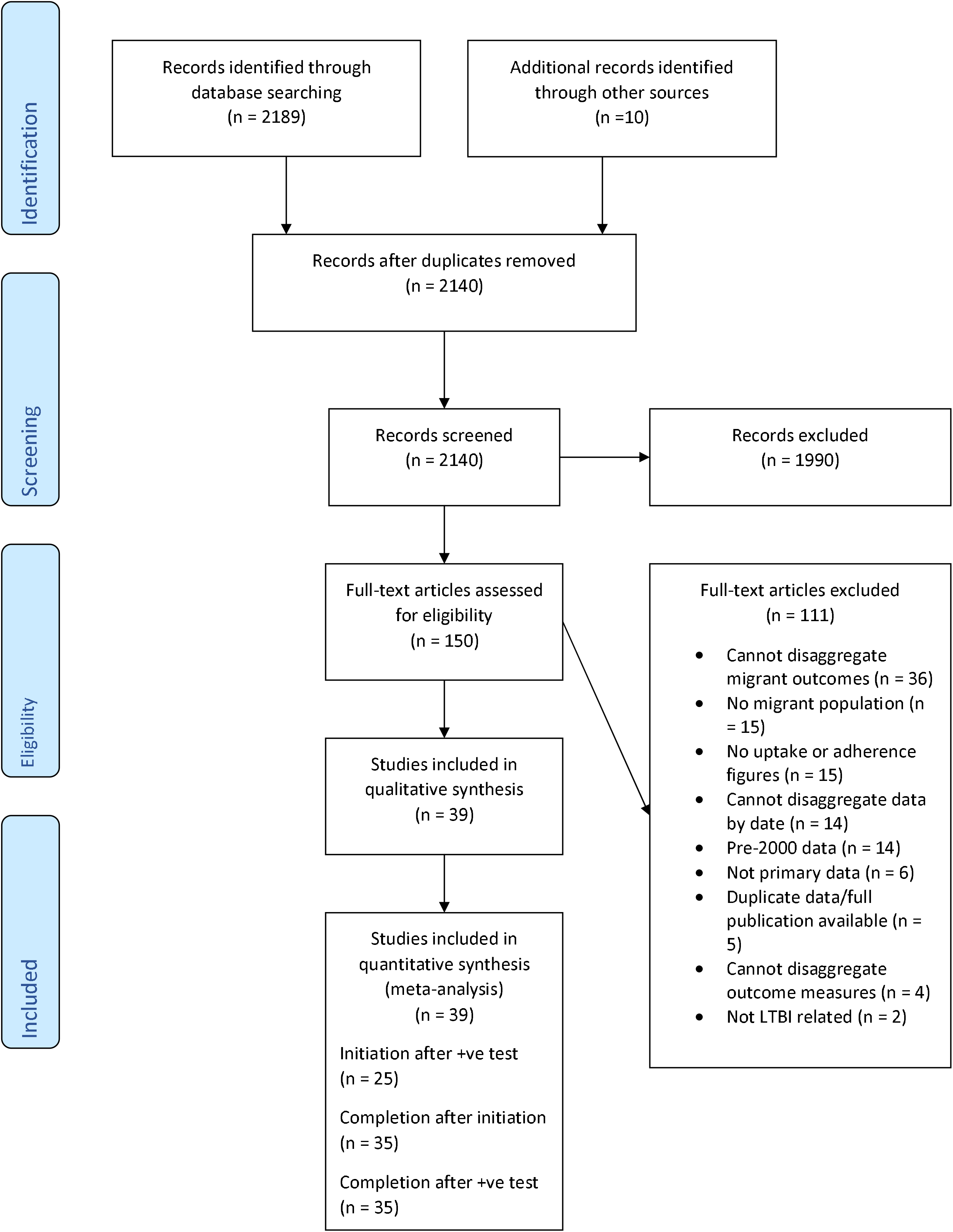
PRISMA flow-diagram of the screening process

### Data analysis

We used a piloted extraction form to retrieve summary estimates relating to initiation and completion, alongside summary data on the study design, dates, and location. We also extracted summaries of analysis relating to positive and negative factors associated with the recorded initiation and completion within each publication.

We carried out statistical analysis within Stata 16.^22^ We used the metapreg module within STATA to calculate the pooled prevalence of treatment initiation, and completion among LTBI positive migrants with corresponding 95% confidence intervals (CIs).^23^ Heterogeneity between studies was examined using the I^2^ statistic. Due to the heterogeneity of the included studies, our analyses utilised random-effects models. We calculated summary estimates of the number of LTBI positive individuals that initiated treatment after testing positive, and the number of LTBI positive individuals who had initiated treatment and completed. We further calculated summary estimates of the full pathway, with regards to the number of LTBI positive individuals that both initiate and complete treatment from publications containing data on both outcomes

We also did a random-effects meta-regression on variables indicated a priori and categorised during the extraction process. Where data were available in the study we categorised each study according to the following (Table 1):

**Table 1.**
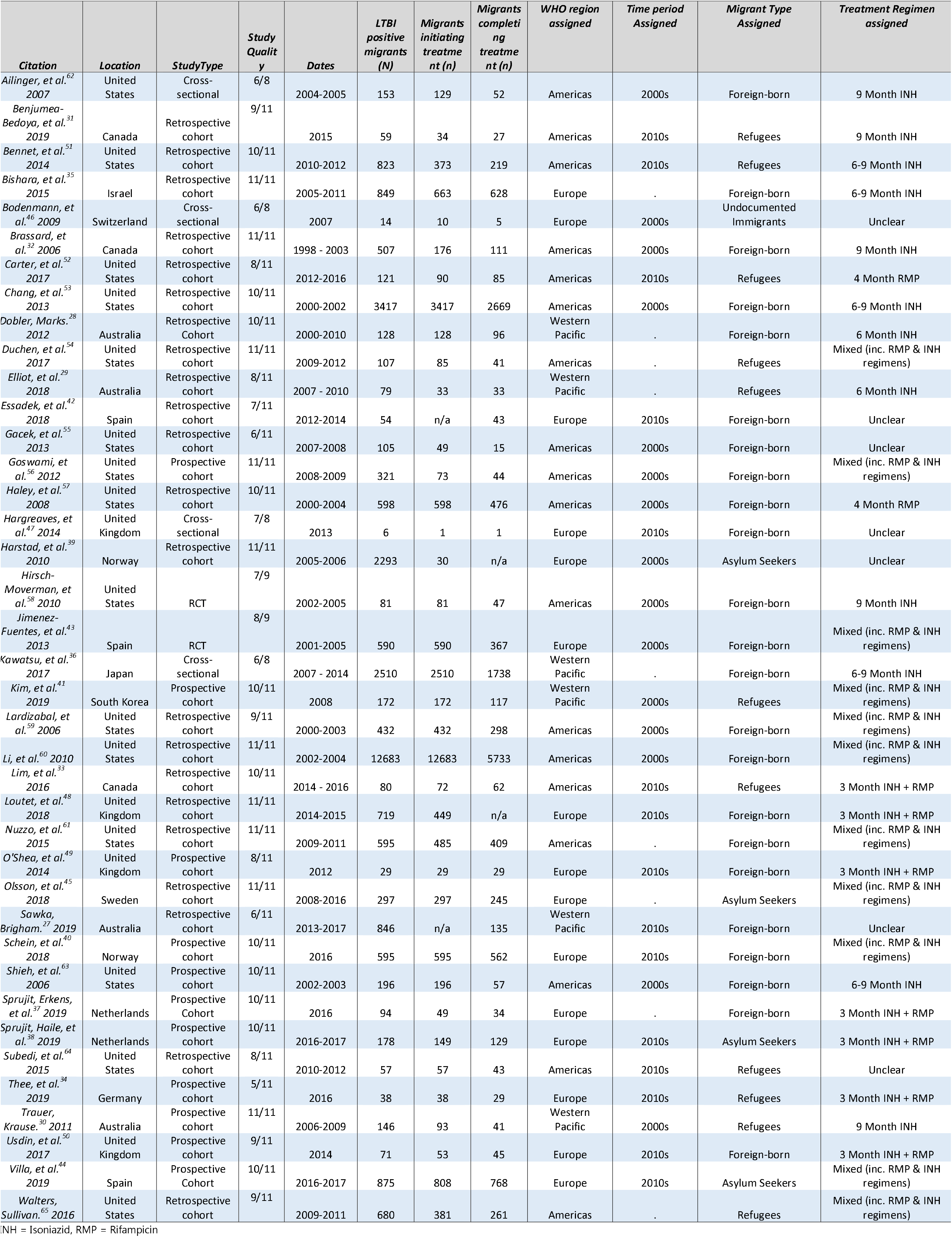
Characteristics of included studies

– The WHO region where the study took place. The decade the data related to (2000 – 2009 or 2010 – 2020).
– The type of migrants being studied (Refugee, asylum-seeker, undocumented, foreign-born).
– The treatment regimens used (6 months INH, 9 months INH, 6-9 months INH, 3 months INH + RMP, 4 Months RMP, mixed (Including RMP & INH regimens), uncertain treatment regimen).
– Treatment setting (A single site for screening & treatment, A single site for treatment only, multiple sites for screening & treatment, multiple sites for treatment only).

These variables were then individually analysed and those resulting in a p-value <0.25 when testing heterogeneity between sub-groups were included in a multivariate model from which results are presented.

Narrative synthesis of positive and negative factors associated with LTBI treatment initiation and completion was also conducted, achieved by utilising statistically significant explanatory variables identified in the included publications’ analyses.

Quality assessment was carried out for each included paper using established appraisal tools. Cross sectional studies were assessed using the Joanna Briggs Institute checklist for prevalence studies.^24^ Cohort studies,^25^ and Randomised Controlled Trials (RCTs),^26^ were assessed using their respective critical appraisal skills programme (CASP) checklists. Using these tools, papers were given a quality score. For case-series and cohort tools, scores were calculated as a total out of the maximum number of applicable questions. Quality scores are reported but studies were not excluded on the basis of quality to increase transparency and ensure all available evidence in this field was reported. Critical appraisal was carried out in duplicate (KR and SHE).

## Results

### Screening results

Database searches yielded 2189 publications, with an additional 10 publications identified through other sources. After removal of duplicates, 2140 studies were subject to title and abstract screening, of those, 1990 citations were excluded. The full texts of 150 publications were screened; 111 records were excluded during full-text screening, and the reasons for their exclusion recorded (Fig 1.). Overall, 39 publications met the inclusion criteria and were included in the review and meta-analysis (Table 1.). The 39 included publications contained data on 31,598 LTBI positive migrants. Studies were done in 13 countries, across three WHO regions (Europe, The Americas, and Western Pacific): Australia (n = 4),^27-30^ Canada (n = 3),^31-33^ Germany (n = 1),^34^ Israel (n = 1),^35^ Japan (n = 1),^36^ The Netherlands (n = 2),^37,38^ Norway (n =2),^39,40^ South Korea (n = 1),^41^ Spain (n = 3),^42-44^ Sweden (n = 1),^45^ Switzerland (n = 1),^46^ The United Kingdom (n = 4),^47-50^ The United States (n = 15).^51-65^

### LTBI initiation after testing positive

In total, 25 studies with outcome data on 8764 migrants were used to calculate the proportion of individuals initiating treatment.^29-35,37-39,44,46-52,54-56,61,62,64,65^ LTBI treatment initiation amongst LTBI positive migrants across the included studies was 69% (95% CI⍰=⍰51–84%; I^2^⍰=⍰99.62%) (Fig 3.). In univariate meta-regression, only the decade in which individual enrolled was found to have a significant effect. Specifically, studies that reported data between in the 2010s) displayed higher initiation (85% initiation; p <0.001) than studies from 2000s (43% initiation; ref.) (Table 2).

**Table 2.**
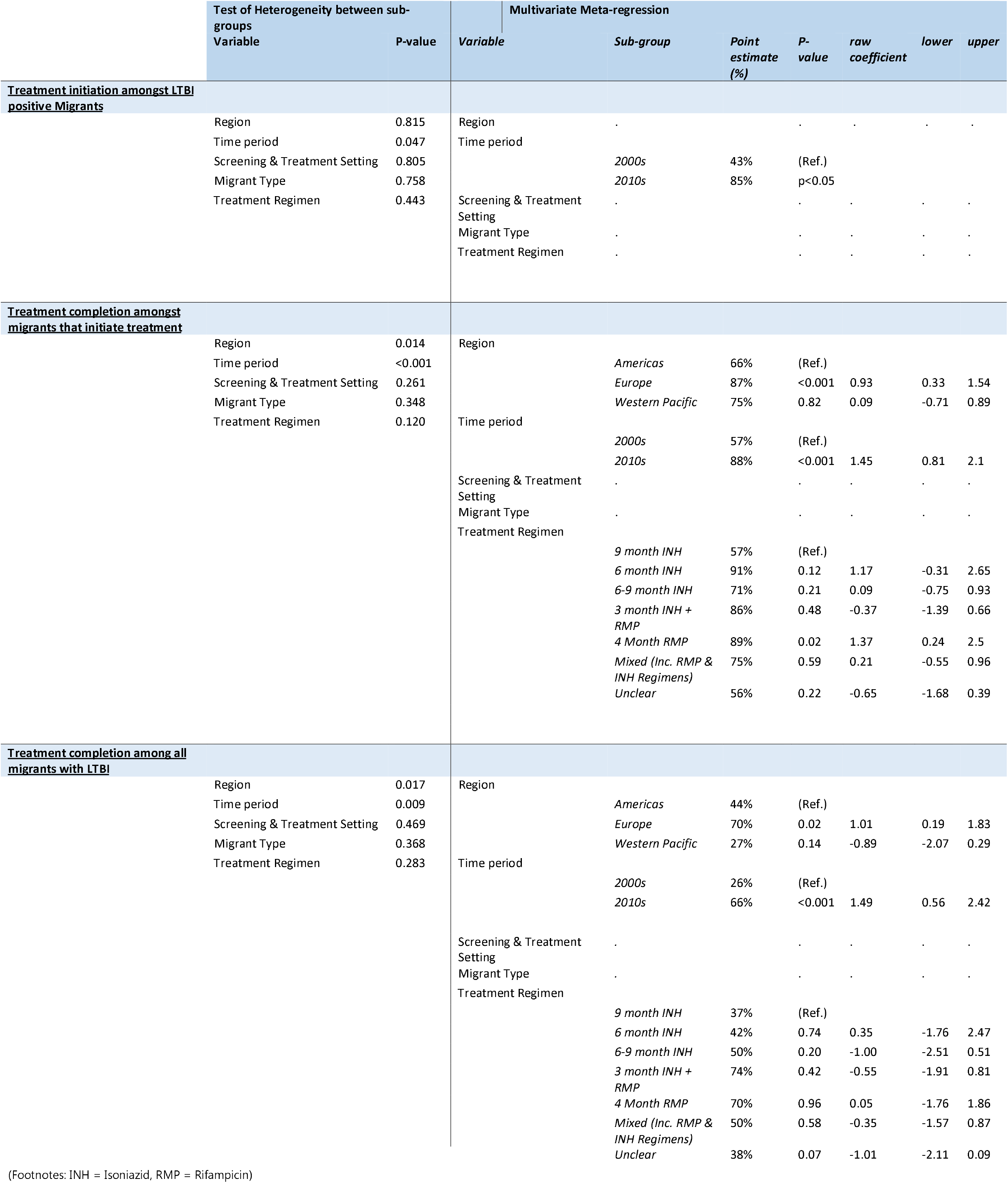
Results of statistical analysis of LTBI treatment initiation and completion outcomes among migrants

Demographic and patient factors associated with non-initiation reported in the studies included (fig 2.): African origin,^51^ being a refugee from sub-Saharan Africa, Northern Africa, or the Middle-East.^65^ Factors positively associated with treatment initiation included having completed 12 or more years of education,^51^ increasing age,^56^ African birthplace,^56^ close contact with an infectious TB case,^56^ non-employment as a precipitating factor in LTBI testing,^56^ having end-stage renal disease.^56^ One study reported significantly higher treatment initiation amongst foreign-born refugees compared to other foreign-born groups and the native-born population.^61^

**Figure 2.**
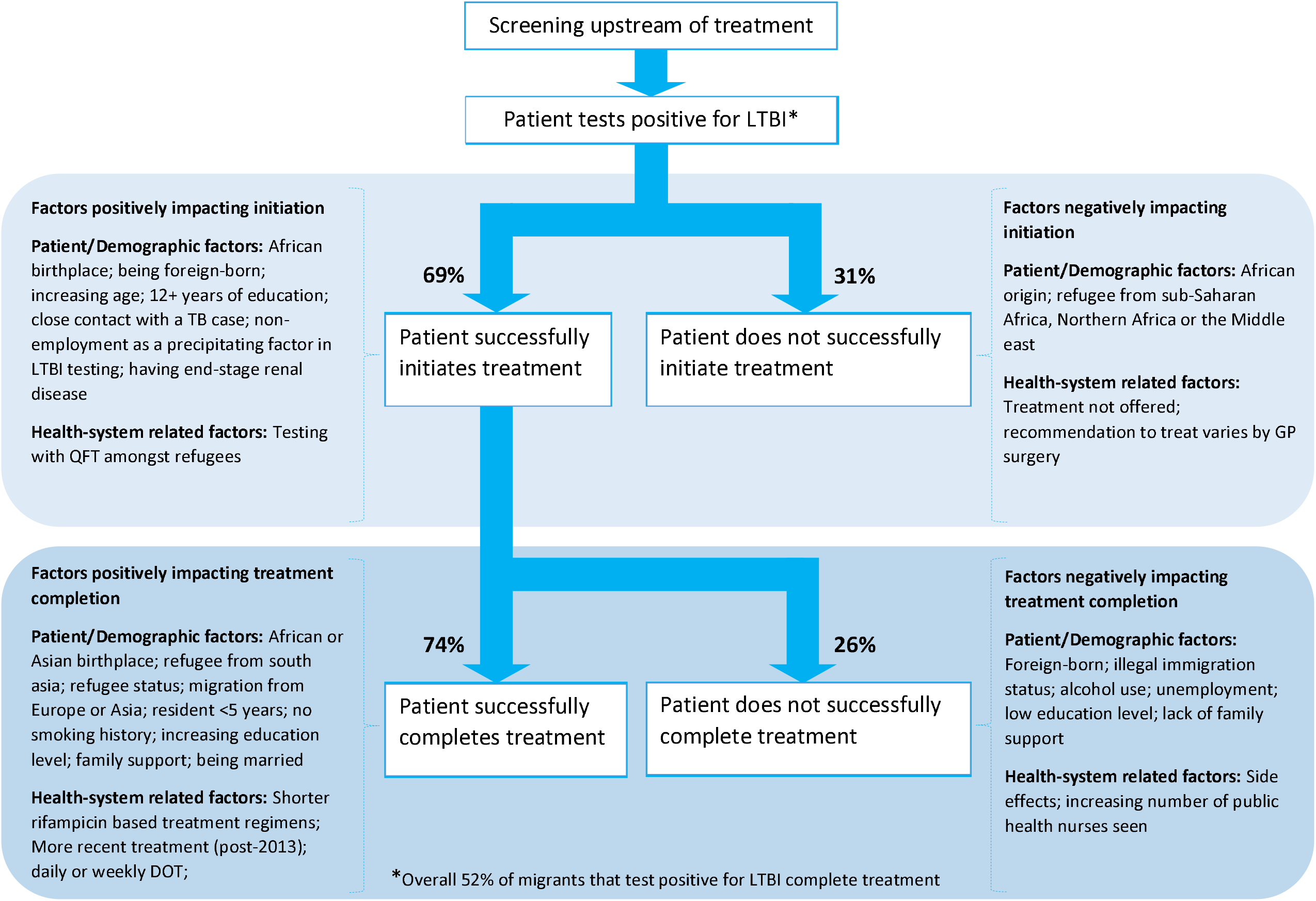
LTBI treatment pathway with associated factors that impact initiation and completion amongst migrants

Reported clinical and systems related factors included (fig 2.): testing with Quantiferon Test (QFT) amongst refugees which led to significantly higher treatment initiation.^65^ However, further studies found non-initiation was associated with treatment not being offered,^55^ and that recommendation to treat varies by primary care clinic.^48^

### LTBI completion after initiation

35 studies with outcome data on 27,451 migrants were used to calculate the proportion of individuals completing treatment subsequent to initiation.^28-38,40,41,43-47,49-65^ LTBI treatment completion amongst migrants initiating treatment was 74% (95% CI⍰=⍰66–81%; I^2^⍰=⍰99.19%) (fig 3.). In univariate meta-regression WHO region, the time period the data related to, and the treatment regimen utilised were all significantly associated with treatment completion. In the multivariate model migrants were more likely to complete treatment in the 2010s (88% completion; p < 0.001) compared to migrants in the 2000s (57% completion; ref.). Migrants treated in the WHO European Region (87% completion; p <0.001) were significantly more likely to complete treatment compared to those in the Americas (66% completion; Ref.) or the Western Pacific (75% completion; p = 0.82). Analysing by treatment regimen, migrants treated with 4 months RMP (89% completion; p = 0.02) were significantly more likely to complete treatment compared to those prescribed 9 months INH (ref.); no other treatment regimen resulted in statistically significant differences (Table 2).

**Figure 3.**
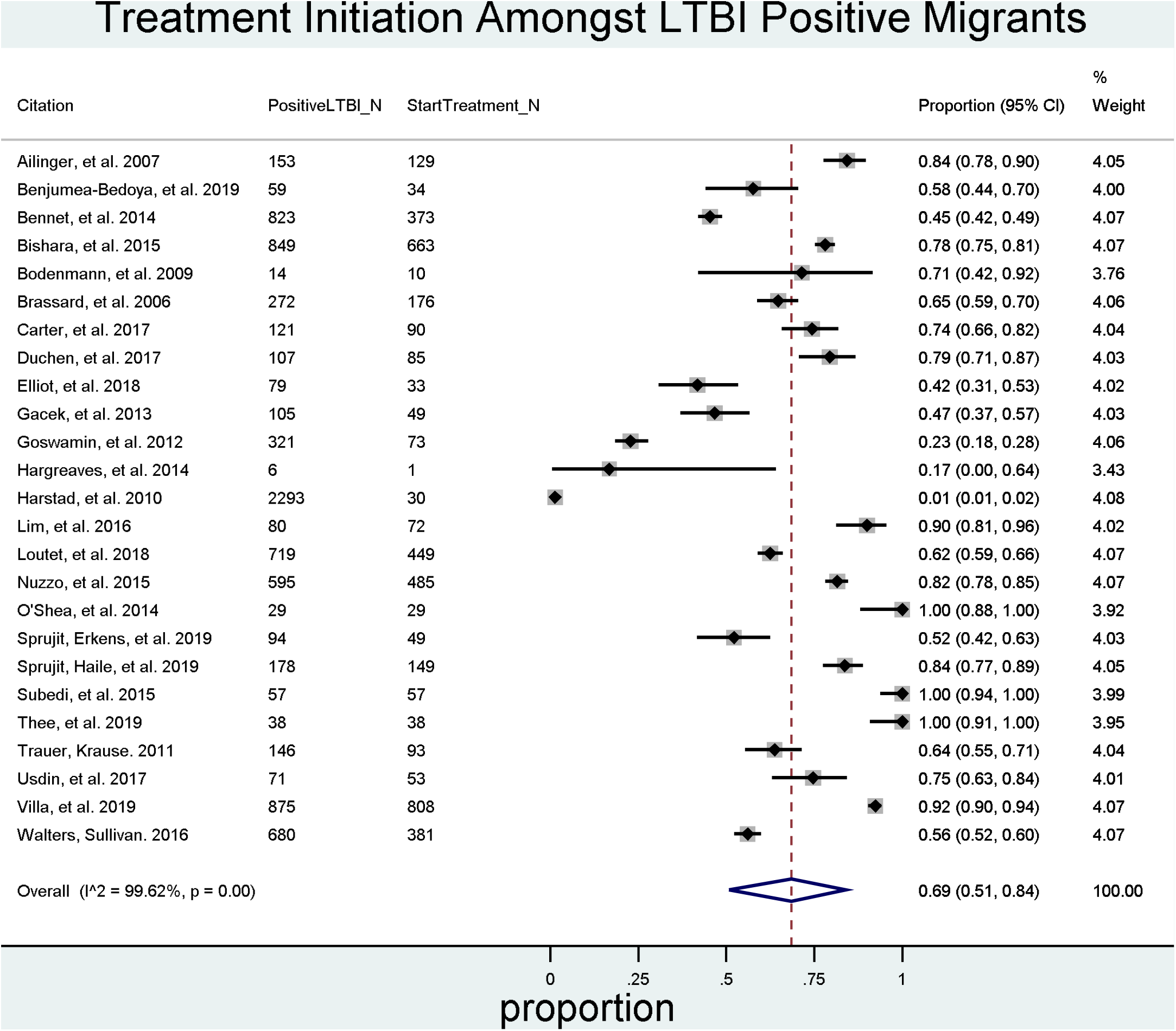
Pooled proportion of LTBI positive migrants that initiate LTBI treatment after testing positive

**Figure 4.**
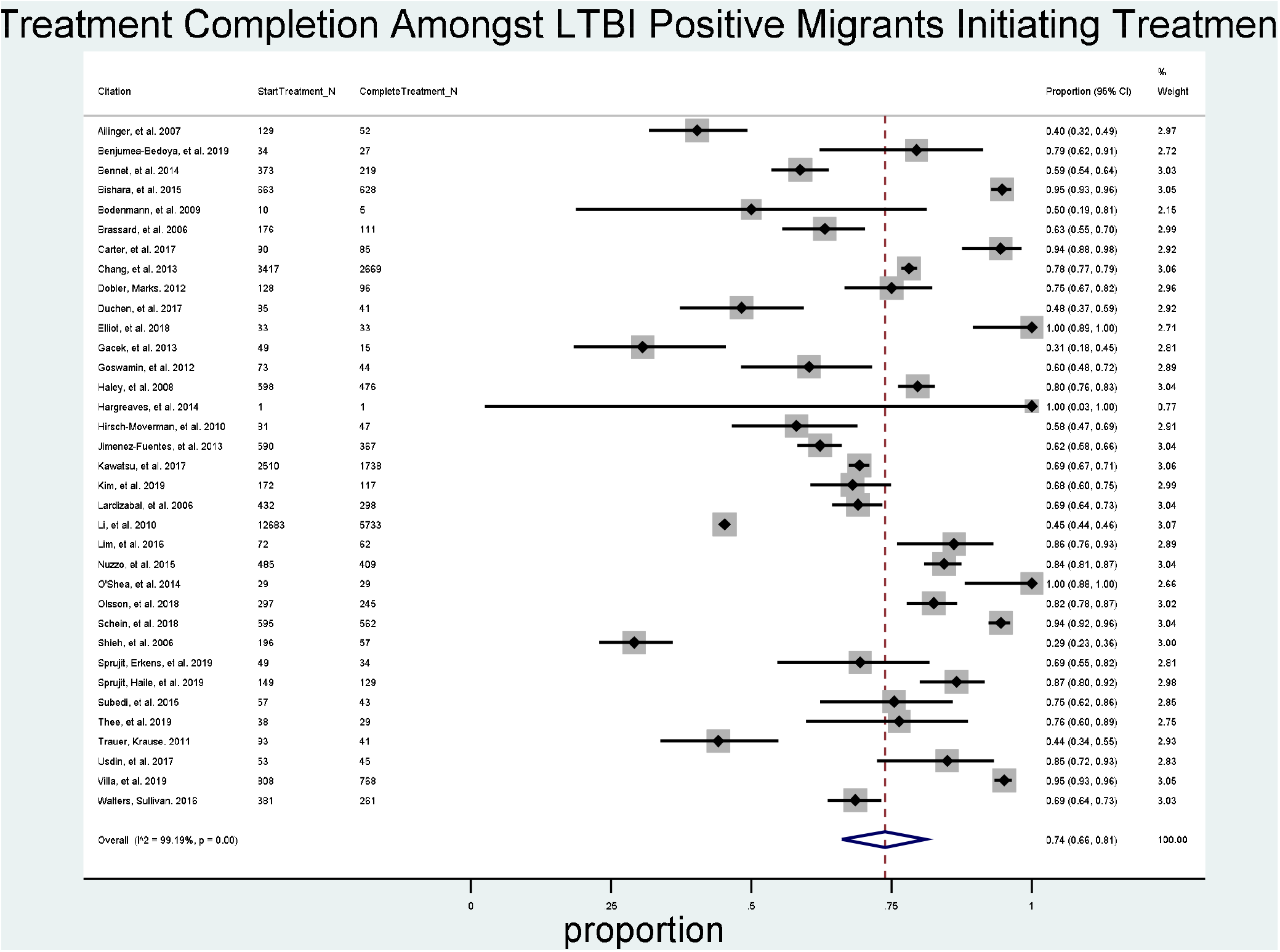
Pooled proportion of migrants that complete LTBI treatment subsequent to initiation

Demographic and patient-related factors associated with non-completion include (fig 2.): foreign-birth,^36,57^ and irregular immigration status.^43^ Alcohol use,^58^ unemployment,^43^ low education level,^43^ and lack of family support,^43^ were also all associated with non-completion of treatment. Demographic and patient-related factors associated with completion included: African or Asian birthplace,^56^ or being a refugee from South Asia.^65^ The absence of tobacco history,^56^ planning to tell friends and family about positive Tuberculin skin test (TST),^56^ prior incarceration,^56^ being married,^58^ and increasing education,^51^ were associated with treatment compliance among foreign-born patients. Foreign-birth,^40,60^ having resided in the host country <5 years,^59^ refugee status^61^ as well as migration from Europe or Asia,^53^ were significantly associated with treatment completion compared to the native-born population within their respective studies. Two studies found no association between any demographic factors and treatment completion in either direction.^62,63^

Clinical and system-related factors associated with non-completion included (fig 2.): side-effects,^28,35,62^ and increasing number of public health nurses seen.^62^ Clinical and system-related factors associated with completion included: In paediatric studies, having two or more family members brought in for screening,^32^ being prescribed a 3RH regimen compared to 6H,^43^ receiving 4R compared to 9H regimen,^59^ 6-month INH vs 9-Month INH,^45^ receiving LTBI treatment in combination with immunosuppressive treatment,^45^ receiving treatment more recently (post-2013),^45^ daily and weekly directly-observed therapy (DOT) was associated with treatment completion among foreign-born patients (vs self-administered care),^40^ and patients taking rifampicin (vs INH) were more likely to complete.^65^

### LTBI treatment completion

25 studies containing outcome data on 6652 migrants were used to calculate the overall proportion of LTBI positive migrants completing treatment.^27,29-35,37,38,42,44,46,47,49-52,54-56,61,62,64,65^ The percentage of migrants completing treatment after screening positive for LTBI was 52% (95% CI⍰=⍰40–64%; I^2^⍰=⍰98.90%) (Fig 5.). In univariate meta-regression WHO region, and time period of treatment receipt were significantly associated with treatment completion. In the multivariate model migrants, were more likely to complete treatment in the 2010s (66% completion; p <0.001) compared to migrants in the 2000s (26% completion; ref.). Migrants treated in the WHO European Region (70% completion; p = 0.02) were also significantly more likely to complete treatment compared to those in the Americas (44% completion; Ref.) and the Western Pacific (27% completion; p = 0.14). No other factors were found to be significantly associated with completion in our analysis (Table 2).

**Figure 5.**
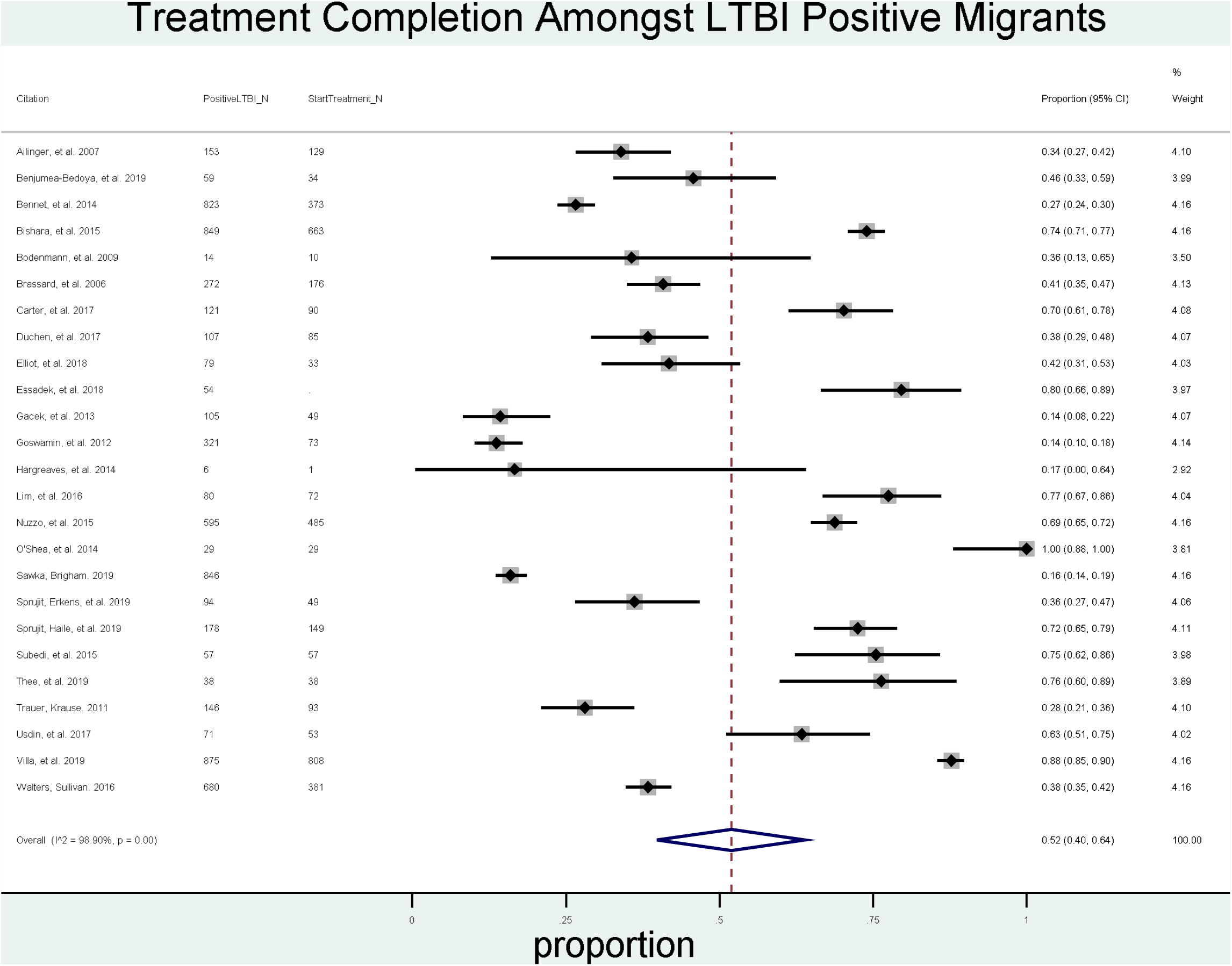
Overall pooled proportion of LTBI positive migrants that initiate and complete treatment.

## Discussion

This is the first systematic review which comprehensively reports on pathway results on migrant LTBI treatment programmes. We identified evidence from 13 mostly high-income low-incidence TB countries, with considerable variation in treatment regimens utilised, and regional differences in treatment outcomes in migrants. Our analysis of 31,598 LTBI positive migrants within the 39 included studies reveals that overall, between 2000 – 2020, 52% of migrants that test positive for LTBI ultimately initiate and complete treatment within studies reporting data on the full pathway. When this was broken down, we found that between 2000 – 2020, 69% of migrants with a positive LTBI test initiated treatment, and 74% of those that initiated treatment, completed it. Overall, these data suggest that there is drop out along the treatment pathway, though this drop out reduced in the latest decade. The included studies highlighted complex barriers and facilitators related to patient demographics and health systems and clinical decision making that impacted on outcomes. Improvements in the LTBI care cascade are needed for migrants, particularly with respect to the initiation of treatment amongst those testing positive. Our data suggest that the current approach requires renewed emphasis and strengthening, despite substantial improvements in outcomes in the past decade.

A previous review of LTBI initiation and completion amongst multiple population groups found initiation rates amongst migrants ranged between 23 – 97%, and completion rates between 7 – 86%.^66^ A further systematic review and meta-analysis including a minority of migrants suggests treatment initiation of 54.6% and completion of 14.3% amongst migrants.^17^ Our study builds upon these estimations by focusing on, and calculating a pooled rate of initiation and completion specifically for migrant populations. In particular our estimation that approximately 74% of migrants that initiate LTBI treatment completed it in the period under study is markedly higher and more optimistic than the 14.3% posited by previous research;^17^ this difference could be attributable to the predominance of pre-2000 data pertaining to the migrant cohorts included in this previous research.

Important socio-demographic and cultural factors may influence the decision of migrant patients to initiate and complete treatment. Research shows stigma and misconceptions regarding TB aetiology is prevalent in some communities. LTBI diagnosis can often be misinterpreted as a TB diagnosis, with fears of stigma and social impact reflecting those of TB.^67^ Clinicians also report difficulties in effectively communicating about LTBI with migrant patients and communities.^19^ In our study, we found that a range of factors, associated with patient demographics, treatment providers, and health-systems that influence LTBI treatment initiation and completion. (Fig 2.). Going forward, more research is needed to better explore and assess facilitators to improve outcomes for LTBI in migrants. Crucially this research must better delineate what it is about being a migrant, or their experiences, that impact outcomes; this could be achieved through greater involvement of migrant communities across the entire research process, from inception to dissemination.

Clinical practice and guidance are also important in ensuring migrants initiate treatment. Our study highlights examples of differing practices, resulting in migrants not initiating treatment due to factors beyond their control.^48,55,65^ Deviation from national guidelines as to screening and treating individuals who are LTBI positive has been evidenced in other research, for example clinicians thinking certain migrants are not eligible for preventative treatment, discrepancies around age thresholds for treatment, and other exclusion criteria.^19,68^. In their recent guidelines, the ECDC call for recently arrived migrants from high-TB-incidence countries to be screened for LTBI using TST or IGRA and for these individuals to be linked to care and treatment;^69^ toolkits supporting best practice for such guidelines exist.^70^ Adaptation of such a toolkit and wider dissemination within, and between, countries focusing LTBI screening amongst migrants may have the potential to alleviate some of the issues encountered in clinical settings.

We found conflicting evidence as to whether foreign-born status acts as a barrier or facilitator regarding treatment initiation and completion in the included studies. Studies using a binary foreign-born status appear to be most ambiguous, with some stating a positive association, ^40,60^ and some a negative association.^36,57^ Studies utilising a wider pool of categories (e.g. regional or country-specific analyses) produce a more nuanced picture, with divergent risk profiles for initiation and completion between migrant groups. This suggests factors such as an individual’s country of origin, years of residency, and legal status provide important context beyond the scope of foreign-born status alone. Future research taking account of these factors will allow us to identify groups requiring greater support with their treatment. The complex and varied composition of migrant groups may also partly explain the differing initiation and completion estimates seen between the WHO regions, which experience different migration flows.^71^ The interaction between factors associated with a migrants’ origin (cultural practices, medical beliefs) and the environment into which they migrate (access to care, availability of tailored services, resultant socioeconomic status), if disentangled, could help to explain the higher levels of treatment initiation and completion seen, for example, in the WHO European Region.

Side-effects and treatment regimen were also associated with treatment outcomes. Whilst side-effects to isoniazid-preventive therapy are common (more so with increasing age), completion of treatment is generally high.^72^ Furthermore, shorter treatment regimens were often associated with better outcomes amongst the included studies and may aid in reducing drop-out due to side-effects. Evidence shows shorter regimens may be as efficacious as longer treatment regimens, and could be associated with greater compliance and less discontinuation due to hepatotoxicity and other side-effects.^11^ Shorter regimens are now more widely recommended.^11,15^ A recent trial HIV-infected patients demonstrates that 1-month daily rifapentine plus isoniazid treatment regimen was non-inferior compared to 9-months of isoniazid alone, and resulted in significantly more patients completing treatment.^73^ If similar ultra-short course regimens can be prescribed to migrant patients, and other barriers to treatment are overcome, they would have the potential to further increase treatment completion. Whilst our analyses found no evidence of a significant association between treatment regimen and outcome, the type of regimen appears to be associated with the time-period under study with more shorter and RMP-containing regimens evidenced in more recent studies. The influence of treatment regimen should not be discounted because of our analysis, with individual-level patient data meta-analysis likely to be better able to analyse this association in the future.

Our analysis is limited by the availability of data available within the published literature. We found conflicting accounts of some explanatory variables (such as being foreign-born) being both positively and negatively associated with initiation and completion. Whilst study level categorisations of treatment regimen were not consistently positively associated with completion, some were approaching significance and future analysis would likely benefit from individual level data. Further granularity of variables, going beyond binary classifications such as being foreign-born are needed, to build up a better picture of the issues in diverse migrant groups (such as refugees, labour migrants, undocumented migrants, students etc). It will also be important to also consider wider socio-economic factors given that we have shown there is still substantial heterogeneity in outcomes that cannot be fully explained by stratifying by migrant type. Analysis of national level programmatic data where possible could also provide a complementary and comparative analysis of treatment initiation and completion.

## Conclusions

Our analysis indicates that there is drop out in the LTBI pathway between screening positive and completing treatment amongst migrants, with multiple complex barriers and facilitators related to patient demographics and health systems and clinical decision making that impacted on outcomes. Improvements in the LTBI care cascade are needed for migrant patients, particularly with respect to the initiation of treatment amongst those testing positive. Our analysis suggests the current approach requires renewed emphasis and strengthening, despite the encouraging improvements in outcomes in the past decade. More meaningful engagement with migrant communities in which they are involved in research may result in more effective design and targeting of interventions and guidelines to support migrant individuals to successfully initiate and complete LTBI treatment. Such steps will address the urgent need to improve the health of these groups alongside meeting International elimination targets for TB.

## Data Availability

The full protocol can be retrieved from PROSPERO (CRD42019140338). Materials including detailed critical appraisal and characteristics of each study are available in the appendices. The extracted data used within our analysis is available from the St Georges University data repository (10.24376/rd.sgul.13521764).

https://doi.org/10.24376/rd.sgul.13521764.v1

## Contributors

SH had the idea for the study and KR, LN, and JL were responsible for the study inception and protocol design. Database searches and screening were done by KR, LN, JL. Data extraction and analysis were done by KR, LN, SH, JL & SEH. Interpretation of the results and drafting of the manuscript was done by KR and SH, with input from all the authors. All authors also commented on and approved the final manuscript.

## Acknowledgments

We would like to acknowledge the experts in the field who we contacted for grey literature and who contributed to the identification and screening of papers and data: Professor Richard Chaisson; Associate Professor Justin Denholm; Professor Stephen Graham; Dr Alexandra Kruse; Dr Tanya Melillo; Dr Drew Posey; Associate Professor Pernille Ravn; Dr Gerard de Vries.

Kieran Rustage is funded by a Rosetrees Trust (PhD studentship grant M775). This study was funded by the European Society of Clinical Microbiology and Infectious Diseases (ESCMID) through a joint ESCMID Study Group for Infections in Travellers and Migrants (ESGITM) and ESCMID Study Group for Mycobacterial Infections (ESGMYC) Research grant. SH is funded by the NIHR (NIHR Advanced Fellowship NIHR300072) and the Academy of Medical Sciences (SBF005\1111). DG is a professor at Saint Camillus International University of Health and Medical Sciences in Rome. SEH is supported by a Medical Research Council PhD studentship (MR/N013638/1).

