## Appendix 1. for "Initiation and completion of treatment for latent tuberculosis infection in migrants globally: A systematic review and meta-analysis"

**Appendix 1.** Representative example of the search strategy (with subject headings) used in this systematic review and meta-analysis (Embase)

1. migrant/ or migrant worker/ or Migrant*.mp.

2. Migrat*.mp.

3.refugee/ or refugee*.mp.

4.asylum seeker/ or asylum seeker*.mp.

5.foreigner*.mp. or foreign worker/

6.foreign born.mp.

7.immigrant/ or immigra*.mp.

8. Emigrants/ or emigrant/ or emigration/ or emigra*.mp.

9. oversea*.mp.

10. foreign student*.mp. or foreign student/

11. international student*.mp.

12. traffick*.mp.

13. 1 or 2 or 3 or 4 or 5 or 6 or 7 or 8 or 9 or 10 or 11 or 12

14. adher*.mp.

15. complian*.mp.

16. deafult.mp.

17. concordan*.mp.

18. treatment outcome/ or treatment outcome*.mp.

19. non adher*.mp.

20. non complian*.mp. or patient compliance/

21. treatment uptake.mp.

22. treatment start.mp.

23. treatment initiation.mp.

24. drop out.mp.

25. follow up/ or loss to follow-up.mp.

26. treatment deferral.mp.

27. treatment completion.mp.

28. treatment success.mp. or treatment outcome/

29. 14 or 15 or 16 or 17 or 18 or 19 or 20 or 21 or 22 or 23 or 24 or 25 or 26 or 27 or 28

30. latent tuberculosis/ or Latent tuberculosis infection.mp.

31. latent TB infection.mp.

32. LTBI.mp.

33. latent tuberculosis.mp.

34. latent TB.mp.

35. latent mycobacterium tuberculosis.mp.

36. inactive tuberculosis infection.mp.

37. inactive tuberculosis.mp.

38.inactive TB.mp.

39. inactive mycobacterium tuberculosis.mp

40. (prophyla* adj3 tuberculosis).mp. [mp=title, abstract, heading word, drug trade name, original title, device manufacturer, drug manufacturer, device trade name, keyword, floating subheading word, candidate term word]

41. 30 or 31 or 32 or 33 or 34 or 35 or 36 or 37 or 38 or 39 or 40

42. 13 and 29 and 41
